# An updated prospective quantitative analysis of symptoms and safety in low-intensity focused ultrasound neuromodulation

**DOI:** 10.64898/2026.06.25.26356569

**Authors:** Aditya Kapoor, Thomas Crahan, Wynn Legon

**Author notes:** Correspondence: Wynn Legon,; 1 Riverside Circle Roanoke, VA, 24016, USA.

## Abstract

Low-intensity focused ultrasound (LIFU) is a non-invasive neuromodulation technique with a favorable safety profile in healthy volunteers. Participant-experienced symptoms however remain inconsistently measured, and prospective benchmarks are lacking. Here, we prospectively characterized symptoms associated with LIFU neuromodulation across eight studies using a standardized Report of Symptoms (ROS). We compiled 629 sessions (472 LIFU, 157 sham) in 106 healthy adults (28.1 ± 9.8 years) across eight cortical and subcortical targets (500 kHz; extracranial ISPPA 3.9–33.3 W/cm²; mechanical index 0.5–1.4). The ROS rated 17 symptom domains from 0 (absent) to 3 (severe) before and after each session. New-onset incidence, symptom severity, and total symptom burden were compared between LIFU and sham. The same instrument was applied in 35 patients with chronic pain. Symptom profiles after LIFU were indistinguishable from sham across all 17 domains. Total symptom burden averaged approximately one domain per session and did not increase after LIFU (0.94 to 1.03; p = 0.120). Post-intervention burden was predicted by baseline burden (β = 0.347, p < 0.001) but not by stimulation condition (p = 0.222). New-onset symptoms did not increase across up to 27 LIFU sessions (OR = 0.99, p = 0.73) and were weakly, non-significantly related to acoustic intensity (ρ = 0.37). Across a prospective, sham-controlled dataset, LIFU added no measurable symptom burden and was well tolerated in healthy adults, with comparable tolerability in patients. These findings establish a benchmark for the safety of human LIFU neuromodulation and a foundation for its therapeutic translation.

## INTRODUCTION

Low-intensity focused ultrasound (LIFU) is a non-invasive neuromodulation technique that transmits acoustic pressure waves through the intact skull to reversibly modulate neuronal activity with millimeter-scale spatial resolution, enabling targeting of superficial and deep brain structures [1,2]. LIFU operates within exposure regimes in which biological effects are thought to arise primarily through mechanical, non-cavitational interactions with tissue rather than irreversible thermal damage. Acoustic exposure in LIFU is characterized using several complementary metrics including acoustic pressure (typically expressed in kilopascals (kPa) or megapascals (MPa), acoustic intensity measures such as spatial-peak pulse-average intensity (I_SPPA_) and spatial-peak temporal-average intensity (I_SPTA_), and the mechanical index (MI), which estimates the likelihood of cavitation-related bioeffects [2].

Early human LIFU neuromodulation studies largely adopted exposure recommendations derived from United States Food and Drug Administration (FDA) diagnostic ultrasound guidelines and the IEC 60601 part 2 standard for therapeutic equipment [3], including limits on I_SPPA_, I_SPTA_, and MI that were originally developed for obstetric and adult cephalic imaging applications. More recently, the International Transcranial Ultrasonic Stimulation Safety and Standards consortium (ITRUSST) proposed consensus recommendations specifically for low-intensity applications of transcranial ultrasound stimulation (TUS), emphasizing both mechanical and thermal safety considerations relevant to neuromodulation paradigms [4]. These recommendations define non-significant biophysical risk exposures as those remaining below thresholds such as MI or transcranial MI (MItc) ≤ 1.9, while also incorporating limits on tissue heating, thermal dose, and exposure duration.

Tissue effects of LIFU have been investigated extensively in both small [5] and large animal models [6,7]. Early work by Lee et al. in sheep reported the presence of small microhemorrhages following highly repetitive sonication of visual cortex at relatively high exposure levels, raising initial concerns regarding potential tissue injury from repeated transcranial ultrasound exposure [6]. However, subsequent well-controlled histological studies in ovine and non-human primate models failed to replicate these findings [7]. In rhesus macaques and sheep, Gaur et al. demonstrated no pre-mortem histologic abnormalities, no evidence of necrosis, apoptosis, edema, inflammation, or hemorrhage, and showed that the sparse extravascular red blood cells occasionally observed were equally present in sham animals and were most consistent with post-mortem extraction artifact rather than ultrasound-induced injury. Importantly, these studies included repeated neuromodulation and MR-acoustic radiation force imaging (MR-ARFI) exposures across a broad range of intensities, including levels exceeding those commonly used in human neuromodulation experiments [7].

In a unique translational opportunity, Stern et al. delivered LIFU in vivo to the anterior temporal lobe of patients with medically refractory temporal lobe epilepsy at least one day before planned resection, targeting the tissue scheduled for removal so that the sonicated tissue could be examined after surgical removal [8]. This examination revealed no detectable damage attributable to ultrasound in seven of eight patients, and neuropsychological testing of the same patients before and after sonication showed no meaningful cognitive decline [8].

Several reviews of human low-intensity transcranial ultrasound stimulation studies have supported a favorable safety profile for neuromodulation within currently employed exposure parameters. In one of the earliest comprehensive reviews, Blackmore et al. concluded that the acoustic parameters used in human neuromodulation studies generally fell within accepted diagnostic ultrasound safety limits and that the available literature supported ultrasound neuromodulation as a safe, non-invasive brain stimulation modality [9].

Subsequently, Pasquinelli et al. systematically reviewed both animal and human focused ultrasound studies and similarly concluded that adverse effects in human studies were rare, with most studies reporting no evidence of clinically meaningful adverse events or tissue injury when LIFU was applied within established neuromodulation exposure regimes [10]. More recently, Sarica et al. reviewed 35 human TUS studies involving 677 participants across healthy and clinical populations and reported no severe adverse events. Mild and transient symptoms occurred in only 3.4% of participants and primarily consisted of headache, scalp heating, neck pain, anxiety, sleepiness, muscle twitches, and transient cognitive complaints [11]. Many of these symptom data were derived from our previous retrospective safety analysis of human LIFU neuromodulation studies [12]. In that report, 120 participants enrolled across seven experiments between 2015 and 2017 (I_SPPA_ = 11.56–17.12 W/cm²) were contacted for follow-up, with 64 participants (53%) completing a telephone-administered symptom questionnaire at intervals ranging from the day of experimentation to 22 months post-participation. No serious adverse events were identified, and 7 of 64 respondents endorsed mild-to-moderate symptoms judged as possibly or probably related to LIFU, yielding an overall positive symptom incidence of 4.3% across all possible questionnaire endorsements. Reported symptoms included neck pain, difficulty paying attention, muscle twitches, and anxiety. Importantly, no persistent or delayed symptoms were identified on follow-up. We additionally observed a significant positive correlation between I_SPPA_ and symptom response rate providing preliminary evidence that acoustic exposure may influence tolerability. Although this report established the first participant-level symptom benchmark for human LIFU neuromodulation, several methodological limitations constrained interpretation, including its retrospective design, lack of pre-stimulation symptom baselines, and a modest response rate that may have introduced self-selection bias. In addition, several constituent experiments employed concurrent TMS, making attribution of symptoms specifically to LIFU difficult to disentangle.

The safety of low-intensity applications of transcranial ultrasound stimulation is currently at the forefront given the recent report of a serious adverse event during a clinical trial targeting the nucleus accumbens in substance use disorder using low-frequency focused ultrasound [13]. In that report, Rezai et al. described the development of edema, petechial microhemorrhage, and persistent neurological sequelae following sonication with a 220 kHz system, with post-hoc analysis suggesting the possibility of inertial cavitation-related injury [13]. Importantly, this report prompted substantial discussion within the ultrasound neuromodulation community regarding the distinction between low-, intermediate-, and high-intensity intervention regimes, the interpretation of “low-intensity” exposures, and the necessity of standardized reporting of acoustic parameters and safety monitoring [14]. Subsequent commentaries and open letters emphasized that many currently accepted low-intensity neuromodulation protocols operate at substantially lower estimated in situ pressures and mechanical indices than those employed in the reported case, and reiterated that adherence to recently established ITRUSST recommendations [4,15], including standardized reporting of pressure, intensity, duty cycle, pulse structure, and transcranial mechanical index (MItc), is critical for cross-study comparison, interpretation of risk, and continued safe translation of the field [14,16]. In parallel, these events have also highlighted the importance of systematic and standardized participant-level symptom monitoring. Recent recommendations have therefore proposed structured frameworks for reporting participant-experienced effects and adverse events in low-intensity ultrasound neuromodulation studies to improve transparency, reproducibility, meta-analytic aggregation, and detection of potential dose-response or exposure-related safety signals across laboratories and devices [17].

Here, we report an updated, prospectively collected assessment of participant-experienced symptoms associated with LIFU neuromodulation in healthy adults using a standardized Report of Symptoms (ROS) instrument [12,17] administered before and after both LIFU and sham sessions. The dataset spans 629 intervention sessions (472 LIFU, 157 sham) in 106 different participants across eight separate studies that administered LIFU to cortical or subcortical brain targets. We additionally applied the same instrument in 35 patients with chronic pain conditions (13 with fibromyalgia and 22 with heterogeneous chronic pain) to provide an initial assessment of tolerability in clinical populations. Primary outcomes included session-level symptom incidence, symptom severity, and within-session symptom change relative to pre-intervention baseline. This study was designed to update our previous report [12] and provide a prospective benchmark for the safety and tolerability of human LIFU neuromodulation.

## METHODS

### Participants

All experimental procedures were approved by the Institutional Review Board at Virginia Tech. All participants provided written informed consent prior to participation. Data from 106 volunteer participants from eight experiments conducted between 2022 and 2026 at the Fralin Biomedical Research Institute at Virginia Tech were analyzed. All participants were volunteers who met study-specific inclusion and exclusion criteria, and received financial compensation for participation. Participants ranged in age from 18–61 years (mean 28.1 ± 9.8 years), with 72 females (67.9%) and 34 males (32.1%). The cohort was predominantly White (72.6%), followed by Asian (18.9%) and Black or African American (6.6%) participants. Hispanic or Latino ethnicity was reported by 5.7% of participants.

### Experiments

A total of 629 sessions were included (472 LIFU, 157 sham) across eight cortical and subcortical targets that included the primary motor cortex (M1) [18], left anterior insula (lAI) [19,20], right anterior insula (rAI) [21], left posterior insula (lPI) [20,22], right posterior insula (rPI), dorsal anterior cingulate cortex (dACC) [19,22,23], prefrontal cortex (PFC), and medial temporal lobe (MTL) (**Figure 1A**). Targets without citations reflect experiments not yet published. LIFU was delivered at 500 kHz, and full device, waveform, parameterization, and coupling details are in Supplementary Methods & **Table 1**.

**Figure 1.**
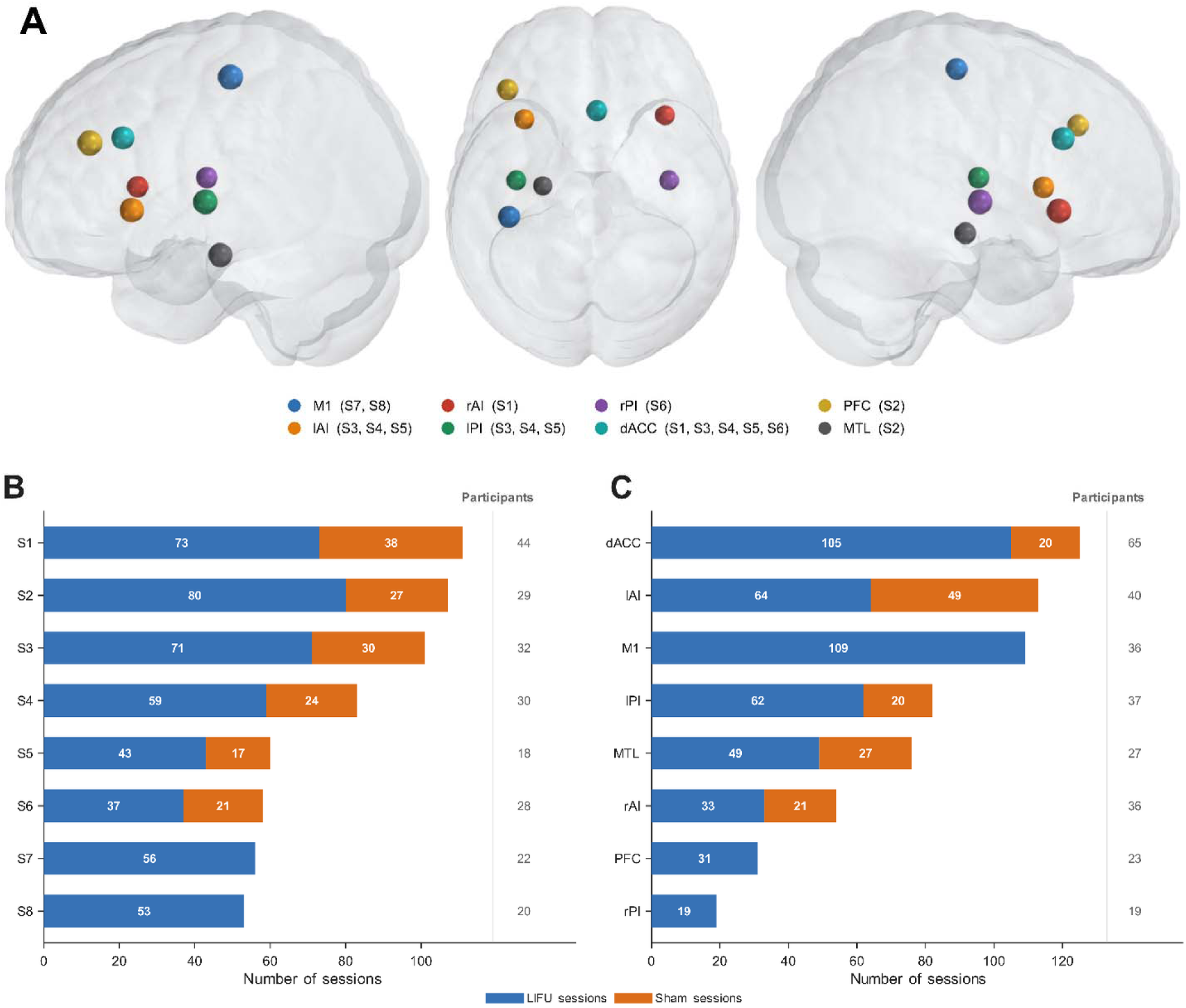
Dataset composition across the eight studies (S1–S8). **A.** The eight studies included LIFU and sham sessions delivered to the primary motor cortex (M1), left and right anterior insula (lAI, rAI), left and right posterior insula (lPI, rPI), dorsal anterior cingulate cortex (dACC), prefrontal cortex (PFC), and medial temporal lobe (MTL), rendered as colored markers on a template brain in left lateral, superior, and right lateral views. **B.** Number of LIFU (blue) and sham (orange) sessions per study, with the number of participants per study shown at right. **C.** Number of LIFU and sham sessions per target, ordered by total sessions, with participants per target shown at right. The dataset comprised 629 sessions (472 LIFU, 157 sham) from a total of 106 participants.

**Table 1.** Acoustic and stimulus parameters by study.

| Study (target) | Fundamental Freq (kHz) | PRF (Hz) | PRI (ms) | Duty Cycle (%) | Pulse Duration (ms) | Pulse Train Duration (ms) | Inter-Stimulus Interval (s) | Total Sonication Duration (s) | Free-Field PNP (kPa) | Isppa (W/cm<sup>2</sup>) | Ispta (W/cm<sup>2</sup>) | MI | Est. Intracranial Pressure (kPa) |
| --- | --- | --- | --- | --- | --- | --- | --- | --- | --- | --- | --- | --- | --- |
| **S1** [21]<br>*rAI, dACC* | 500 | 10 | 100 | 30 | 30 | 40,000 | N/A<br>(Single 40 s block;<br>Inter-pulse off = 70 ms) | 40<br>(Per target) | ~900 | 27 | 8.1 | ~1.27 | NR |
| **S2\***<br>*PFC, MTL* | 500 | 10 | 100 | 30 | 30 | 1,000 | 1 | 96<br>(1 s on / 1 s off) | NR | 6.16 | 1.85 | 0.61 | NR |
| **S3** [19,22,23]<br>*dACC, lAI, lPI* | 500 | 1,000 | 1 | 36 | 0.36 | 1,000 | 5 | 100<br>(100 × 1 s pulses) | 380 (AI/PI)<br>400 (dACC) | 4.2 (AI/PI)<br>4.5 (dACC) | 1.5 (AI/PI)<br>1.62 (dACC) | 0.54 (AI/PI)<br>0.57 (dACC) | AI: 180.1 ± 84.6<br>PI: 262.1 ± 93.6<br>dACC: 119.8 ± 55.7<br>(mean ± SD; N = 16) |
| **S4** [19,22,23]<br>*dACC, lAI, lPI* | 500 | 1,000 | 1 | 36 | 0.36 | 1,000 | 10–20<br>(randomized) | 40<br>(40 × 1 s) | 368.5–400 | 3.5–4.34 | 1.56 | 0.52–0.57 | AI: 179.3 ± 75.1<br>PI: 263.5 ± 93.4<br>dACC: 115.2 ± 53.1<br>(mean ± SD; N = 23) |
| **S5** [20]<br>*dACC, lAI, lPI* | 500 | 1,000 | 1 | 30 | 0.30 | 2,500 | 14–20<br>(jittered) | 50<br>(20 × 2.5 s) | 750 | 18.75 | 5.63 | 1.06 | NR |
| **S6\***<br>*dACC, rPI* | 500 | 10 | 100 | 30 | 30 | 1,000 | N/A | 40<br>(40 × 1 s) | NR | 33.33 | 10 | 1.43 | NR |
| **S7** [18]<br>*M1* | 500 | 1,000 | 1 | 1, 10, 30, 50, 70 | 0.01–0.7 | 100 or 500 | 10 | 2–10<br>(20 trials × 100 or 500 ms) | ~424 (6 W/cm<sup>2</sup>)<br>~888 (24 W/cm<sup>2</sup>) | 6 or 24 | 0.06–16.8 | 0.60–1.26 | 141.1 ± 66.7 (6 W/cm<sup>2</sup>)<br>275.3 ± 130.2 (24 W/cm<sup>2</sup>)<br>(mean ± SD; N = 18) |
| **S8\***<br>*M1* | 500 | 10, 100, 1,000 | 100, 10, 1 | 1, 5, 30 | 0.01–30 | 10,000 | N/A<br>(Single 10 s block) | 10 | 380<br>760 | 6 or 24 | 0.06–7.2 | 0.54<br>1.07 | NR |

Studies are designated S1–S8. Bracketed numbers indicate the manuscript reference citation(s) for each published protocol.
\* Unpublished study; full acoustic characterization and intracranial pressure modeling are pending and reported as NR.
NR, not reported; N/A, not applicable (continuous single-block sonication, no inter-stimulus interval).
Derived quantities: PRI = 1000 / PRF; pulse duration = duty cycle × PRI; *Ispta* = *Isppa* × duty cycle. MI = PNP (MPa) / √f (MHz). Estimated intracranial pressures are simulation-based (mean ± SD across modeled participants).
Abbreviations: PRF, pulse repetition frequency; PRI, pulse repetition interval; PNP, peak negative pressure; *Isppa*, spatial-peak pulse-average intensity; *Ispta*, spatial-peak temporal-average intensity; MI, mechanical index; rAI/lAI, right/left anterior insula; rPI/lPI, right/left posterior insula; dACC, dorsal anterior cingulate cortex; PFC, prefrontal cortex; MTL, medial temporal lobe; M1, primary motor cortex.

### Report of Symptoms

Symptom monitoring was conducted at each study visit using a Report of Symptoms (ROS) assessment [12,17], which assessed the presence and severity of 17 symptom domains on a 4-point ordinal scale: 0 = absent, 1 = mild, 2 = moderate, 3 = severe. Domains assessed were: headache, scalp discomfort, neck pain, tingling, itchiness, sleepiness, attention difficulty, mood change, tooth pain, hearing change, nausea, muscle twitches, dizziness, anxiety, forgetfulness, balance difficulty, and hand sensation. The ROS was administered twice per session, once before the intervention and once at least 30 minutes after it, where intervention denotes LIFU sonication or sham. These paired pre-intervention and post-intervention timepoints allowed within-session comparison of symptom ratings. A symptom domain was considered endorsed at a given timepoint if the participant rated the symptom as mild or greater (ordinal score ≥ 1). The ordinal severity rating of “severe” (score = 3) reflects the participant’s subjective rating of maximal symptom intensity within the ROS framework and does not constitute a regulatory classification of a serious adverse event.

## Data Analysis

### Symptom Endorsement

This analysis addresses whether the overall prevalence of each symptom domain changed from pre- to post-intervention, pooled across all 629 sessions regardless of stimulation condition (LIFU or sham) or target. Session-level endorsement rates were computed for each of the 17 ROS domains at the pre-intervention and post-intervention timepoints across all 629 sessions. Within-session change in endorsement status was evaluated using McNemar’s test for paired proportions [24], applied separately to each domain, with p-values adjusted across all 17 domain-level tests using Benjamini-Hochberg false discovery rate (BH-q) [25]. Net endorsement change was expressed in percentage as the post-intervention rate minus the pre-intervention rate, such that positive values indicate increased endorsement and negative values indicate decreased endorsement post-intervention.

### New onset symptom profile

Where the endorsement analysis captures overall prevalence change, this analysis quantifies symptom incidence: how often a symptom that was absent at pre-intervention newly appeared post-intervention, using only sessions in which that symptom was absent at baseline. A new-onset symptom event was defined as endorsement of a symptom domain at the post-intervention assessment (ordinal score ≥ 1) when that same domain was rated absent at pre-intervention within the same session. Analyses were restricted to sessions considered “at risk” for new onset within a given domain. Specifically, only sessions in which the symptom was absent at pre-intervention contributed to the denominator for that domain. Accordingly, denominators varied across symptom domains depending on baseline symptom prevalence. For each symptom domain, the new-onset rate was calculated as the proportion of at-risk sessions in which the symptom was newly endorsed following intervention. Wilson score 95% confidence intervals were computed for each domain-level estimate [26]. An overall pooled new-onset rate was additionally calculated as the proportion of all sessions in which at least one symptom domain was newly endorsed post-intervention. This pooled estimate served as the reference value for subsequent study-level and brain target-level comparisons.

Symptom domains were additionally categorized according to absolute new-onset frequency as rare (<5%), uncommon (5–15%), or common (>15%). These zones are an operational convention adopted for descriptive presentation in this dataset.

New-onset rates were further estimated separately at the study level and brain target level to characterize variability in participant-experienced symptom profiles across experimental contexts. For each study and target, the rate was defined as the proportion of sessions in which at least one symptom domain was newly endorsed following intervention. Bootstrapped 95% confidence intervals were generated using 5,000 resampling iterations with replacement. The pooled session-level new-onset rate across all 629 sessions served as the reference for these comparisons. Domain-level new-onset rates were compared across brain targets and across studies using a label-permutation χ² test (10,000 permutations), with BH-q correction applied across all 17 domains.

### Symptom Severity Transitions

Whereas the endorsement and new-onset analyses classified each symptom only as present or absent, this analysis evaluated within-session change across all four severity levels (absent, mild, moderate, severe) for each domain. For each of the 17 ROS domains, the paired pre- and post-intervention severity ratings (0–3, absent to severe) were compared with the Wilcoxon signed-rank test, with p-values adjusted across the 17 domains using BH-q. Anxiety and sleepiness were prespecified for detailed severity analysis on the basis of baseline prevalence and within-session change (in reference to the full dataset). Endorsement rates were computed separately for LIFU and sham sessions at the pre- and post-intervention timepoints with Wilson 95% confidence intervals, and the post-intervention endorsement rates were compared between conditions using Fisher’s exact test.

### LIFU versus sham Comparison

This analysis was conducted independently of the paired pre/post endorsement analyses to specifically determine whether LIFU intervention was associated with a differential pattern of newly emerging symptoms relative to sham. New-onset rates were compared between LIFU (n = 472) and sham (n = 157) conditions for each of the 17 ROS domains using Fisher’s exact test, with BH-q correction applied across all comparisons. For each domain, the new-onset rate was calculated using the domain-specific at-risk denominator, defined as sessions in which that symptom was absent at pre-intervention baseline within the corresponding condition. Because domain-level analyses were restricted to at-risk sessions, these rates are not directly comparable to the pooled session-level new-onset rate, which reflects endorsement of any symptom domain within a session. Absolute risk differences (ARDs) were calculated for each symptom domain as the LIFU new-onset rate minus the sham new-onset rate, expressed in percentage. Positive ARD values indicate higher incidence during LIFU, whereas negative values indicate higher incidence during sham. Ninety-five percent confidence intervals for ARDs were computed using the Newcombe hybrid score method [27].

To contextualize the magnitude of between-condition differences independently of statistical significance, absolute risk differences were referenced against a ±5 percentage-point band. Domains whose difference exceeded this band were identified descriptively, and the direction of the difference (LIFU > sham or sham > LIFU) was examined as secondary evidence regarding the specificity of observed symptom patterns to LIFU.

### Total Symptom Burden

Where the preceding domain-level analyses characterized each symptom separately, this analysis reduced the 17 ROS domains to a single per-session measure. Total symptom burden was defined as the number of ROS domains endorsed (rated mild or greater) within a session, an integer score ranging from 0 (no domain endorsed) to 17 (all domains endorsed) that represents how many distinct symptoms a participant reported at a given timepoint. Burden was computed at the pre-intervention and post-intervention assessment of each session, and within-session change was evaluated with the Wilcoxon signed-rank test applied to the paired pre/post scores. This comparison was performed separately for LIFU (n = 472) and sham (n = 157) sessions, and descriptive statistics (mean ± SD, median) were reported for each condition and timepoint. Post-intervention total symptom burden served as the outcome variable in a linear mixed-effects model described in *Predictors of Post-Intervention Symptom Burden* below, linking these session-level distributions to the predictors of post-intervention burden.

### Predictors of Post-Intervention Symptom Burden

A linear mixed-effects model (LME) evaluated whether LIFU or sham predicted post-intervention total symptom burden beyond baseline symptom state. Post-intervention total symptom burden served as the continuous outcome, and two fixed effects were entered: (i) pre-intervention total symptom burden (continuous), to account for each session’s baseline symptom state, and (ii) stimulation condition (LIFU = 1, sham = 0), to test whether LIFU exposure predicted post-intervention burden above baseline. Participant was included as a random intercept to account for the repeated sessions contributed by individual participants. Fixed-effect coefficient estimates (β), standard errors, 95% confidence intervals, and p-values are reported for both predictors.

### Longitudinal Safety Across Repeated LIFU Exposure

As LIFU advances toward therapeutic applications, characterizing safety across repeated sessions is a necessary translational benchmark that single-session safety reports cannot provide. Focused ultrasound at neuromodulatory parameters does not produce cumulative tissue dose effects [4,7,8,28–30]. Repeated non-invasive brain stimulation can nonetheless induce cumulative changes in neural excitability and cortical plasticity across sessions [31,32], and longitudinal symptom surveillance is standard practice in non-invasive brain stimulation safety monitoring [33]. A subset of participants in the present dataset enrolled across multiple studies, accumulating up to 27 LIFU sessions across diverse cortical and subcortical targets, providing within-participant symptom data across an exposure range that extends beyond what prior LIFU safety reports have characterized.

For each participant, LIFU sessions were numbered chronologically to give a within-participant LIFU session number, with the first LIFU session received numbered 1, the second 2, and so on. LIFU and sham sessions were numbered separately so that this value indexed cumulative LIFU exposure rather than total study visits. Sessions were grouped into eight within-participant session-number bins (1, 2, 3, 4, 5, 6–8, 9–12, and ≥13), with single-session bins for the first five LIFU sessions and progressively wider bins thereafter to retain a sufficient number of sessions per bin for stable rate estimation. For each bin, the new-onset rate was computed as the proportion of at-risk sessions in which at least one ROS domain was newly endorsed at post-intervention, and a 95% confidence interval was obtained by resampling the at-risk sessions in that bin with replacement across 5,000 iterations. To test whether cumulative LIFU exposure predicted new-onset symptoms, a generalized linear mixed-effects model (GLME) with a logit link was fit to the session-level data, with new-onset status as the binary outcome, within-participant LIFU session number as a continuous fixed-effect predictor, and participant as a random intercept to account for repeated sessions within individuals. Fixed-effect coefficient estimates (β), standard errors, odds ratios (OR = exp(β)), and p-values are reported.

### Acoustic Intensity and Symptom Incidence

Whereas the preceding analyses characterized symptom incidence within and between conditions, this analysis tested whether the intensity of intervention related to symptom incidence across studies, updating the study-level association reported previously [12]. The unit of analysis was the study, and the outcome was the new-onset rate, defined as the proportion of sessions in which at least one of the 17 ROS domains was newly endorsed at post-intervention, computed separately for LIFU and sham sessions within each study. Each study’s extracranial (free-water) ISPPA was taken as the mean across that study’s sessions (**Table 1**). Six studies that included a sham condition were analyzed (S1, S2, S3, S4, S5, and S6, totaling 363 LIFU and 157 sham sessions). Two motor cortex studies were excluded because they delivered more than one intensity that could not be attributed to individual sessions and contained no sham sessions (S7 and S8). The association between extracranial ISPPA and the new-onset rate was quantified with the Spearman rank correlation for LIFU sessions, and sham sessions were assigned their own study’s extracranial ISPPA and analyzed identically as a negative control. Sensitivity to individual studies was assessed by recomputing the coefficient with each study removed in turn. Per-study rates were summarized with Wilson 95% confidence intervals, and the relationship was summarized with ordinary least squares fit and its 95% confidence interval.

### Symptom Safety in Clinical Pain Populations

To examine whether the within-session symptom profile observed in healthy adults extends to patient populations, the same ROS assessment was applied in two independent clinical cohorts comprising 35 patients in total that were not part of the eight healthy-adult studies and contributed no sessions to any preceding analysis. Cohort 1 comprised 13 participants with fibromyalgia, and Cohort 2 comprised 22 participants with heterogenous chronic pain. Both cohorts received LIFU and sham intervention in within-subjects designs, and the ROS was administered before and after each session as in the main dataset. Full acoustic parameters, demographics, and procedures for the fibromyalgia cohort are reported separately [34], and the chronic pain cohort is unpublished, so only its symptom data are presented here. For each cohort, total symptom burden, defined as the number of ROS domains endorsed within a session, was computed at the pre- and post-intervention timepoints, and within-session change was evaluated with the Wilcoxon signed-rank test applied separately to LIFU and sham sessions.

A significance threshold of α = 0.05 was applied to all statistical tests. Where multiple comparisons were performed across related outcomes, p-values were adjusted using BH-q. Descriptive statistics are presented as mean ± SD unless otherwise specified.

## RESULTS

### Dataset Characteristics

The compiled dataset comprised 629 sessions from 106 participants across eight independent studies conducted between March 2022 and February 2026 at the Fralin Biomedical Research Institute at Virginia Tech. To provide neutral descriptors and avoid naming unpublished protocols, the studies are designated S1 to S8, ordered by session count, and together delivered LIFU to eight cortical and subcortical targets shown on the brain in **Figure 1A**. S1 (n = 111) targeted the right anterior insula (rAI) and dorsal anterior cingulate cortex (dACC) [21]. S2 (n = 107) targeted the prefrontal cortex (PFC) and medial temporal lobe (MTL). S3 (n = 101) and S4 (n = 83) each targeted the dACC, left anterior insula (lAI), and left posterior insula (lPI) [19,22,23]. S5 (n = 60) targeted the dACC, lAI, and lPI [20]. S6 (n = 58) targeted the dACC and right posterior insula (rPI). S7 (n = 56) and S8 (n = 53) targeted the primary motor cortex (M1), with S7 reported previously [18]. Of the 629 sessions, 472 (75.0%) were LIFU and 157 (25.0%) were sham (**Figure 1B**), and the number of LIFU and sham sessions delivered to each target is shown in **Figure 1C**. The imbalance between LIFU and sham session counts reflects study design, as protocols targeting multiple regions contributed one LIFU session per target per participant but a single sham session irrespective of the number of targets.

Reflecting the multi-study structure of the dataset, 56 of 106 participants (52.8%) enrolled in more than one study (median 2 studies, range 1–7), contributing repeated sessions across different targets and protocols, with per-participant session (LIFU and sham) counts ranging from 1 to 32 (mean 5.9, median 4).

### Symptom Endorsement

Symptom endorsement rates, defined as the proportion of sessions in which a given domain was rated mild or greater, were computed for all 17 ROS domains at both pre- and post-intervention across the 629 sessions. Rates were computed across all sessions regardless of LIFU or sham, as pre-intervention symptom burden reflects participant state prior to any intervention. The most commonly endorsed pre-intervention symptoms were sleepiness (40.4%), anxiety (22.7%), attention difficulty (9.9%), and neck pain (5.6%) (**Figure 2A**). The remaining 13 domains were each endorsed in fewer than 5% of sessions.

**Figure 2.**
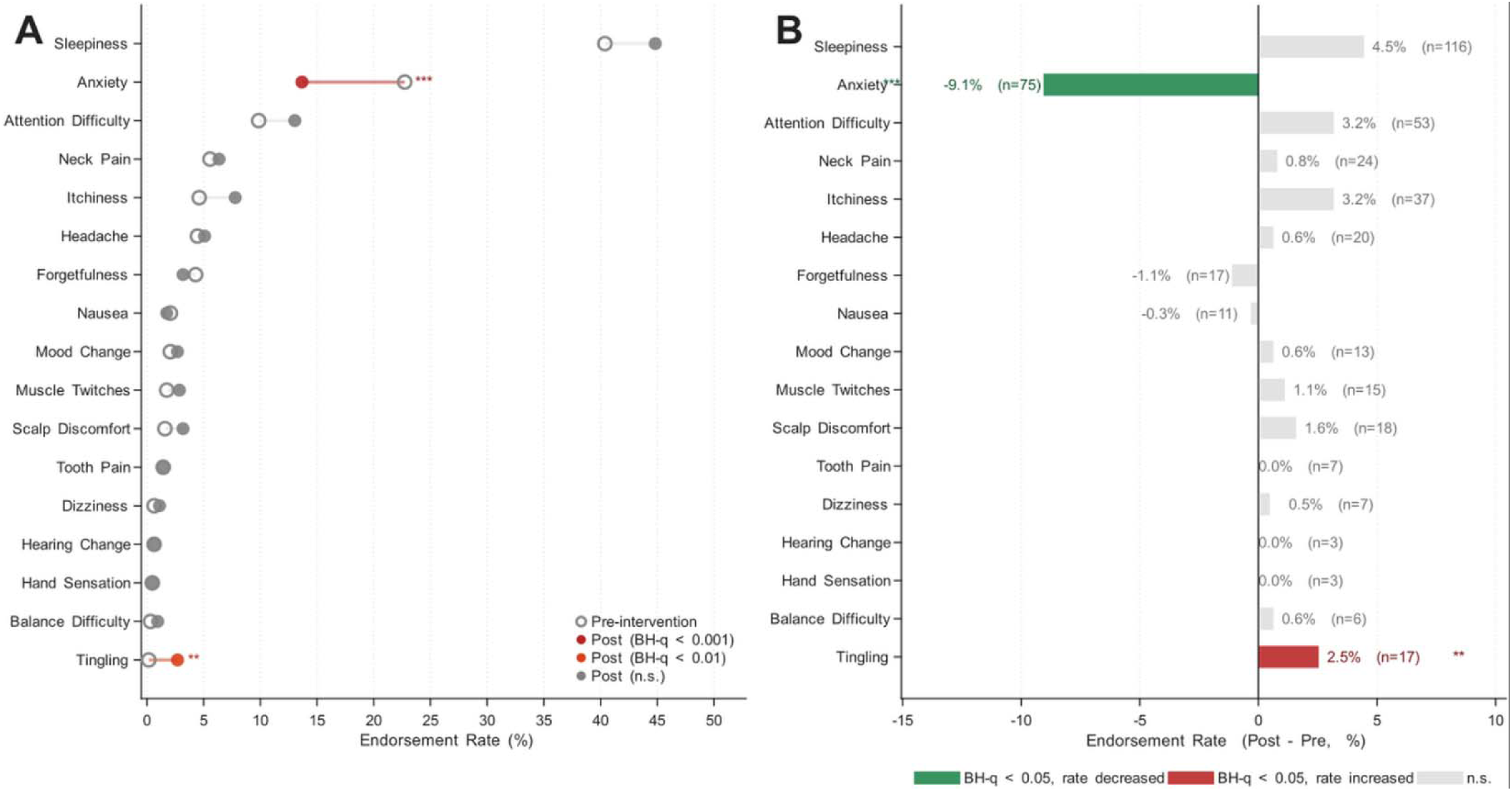
Pre- and post-intervention symptom endorsement across all 629 sessions (LIFU and sham). Endorsement was defined as a rating of mild or greater on the 4-point ordinal scale (absent, mild, moderate, severe). **A.** Pre-intervention (open circles) and post-intervention (filled circles) endorsement rates for all 17 symptom domains, ordered by ascending pre-intervention rate. Connecting lines indicate direction of change. Filled circles are color-coded by BH-q significance tier (McNemar test, BH-q correction, n = 629 sessions). Significance markers (** BH-q < 0.01, *** BH-q < 0.001) appear adjacent to whichever of the pre- or post-intervention rates is higher for that domain. **B.** Net change in endorsement rate (post minus pre, percentage) for each domain. Bars extending left indicate decreased post-intervention endorsement (green, BH-q < 0.05). Bars extending right indicate increased endorsement (red, BH-q < 0.05). Gray bars indicate non-significant change. Each bar is annotated with the net change in percentage and the count of sessions that shifted in that direction (endorsed-to-absent for decreasing domains, absent-to-endorsed for increasing domains), out of all 629 sessions.

Two domains reached BH-q significance. Anxiety endorsement decreased from 22.7% pre-intervention to 13.7% post-intervention (McNemar χ² = 33.7, BH-q = 1.08 × 10⁻□). Tingling showed a significant net increase from 0.2% pre-intervention to 2.7% post-intervention (McNemar χ² = 12.5, BH-q = 0.003). Across the 17 domains, net endorsement change ranged from −9.1% (anxiety) to +2.5% (tingling) (**Figure 2B**).

### New-Onset Symptom Profile

New-onset events were defined as endorsement of a domain at the post-intervention assessment among sessions in which that domain was rated absent at the pre-intervention baseline. Across all 629 sessions, 237 of 629 (37.7%) had at least one domain newly endorsed post-intervention, comprising 35.4% of the 472 LIFU sessions and 44.6% of the 157 sham sessions. Across domains, new-onset rates spanned all three frequency zones (rare, uncommon, and common). Sleepiness was the only domain in the common zone and the most frequently newly endorsed symptom (30.9%). Itchiness (6.2%) and attention difficulty (9.3%) fell in the uncommon zone, and the remaining 14 domains were rare. The two domains with a significant overall change in endorsement, anxiety and tingling, had low new-onset rates of 3.7% and 2.7%, respectively (**Figure 3A**).

**Figure 3.**
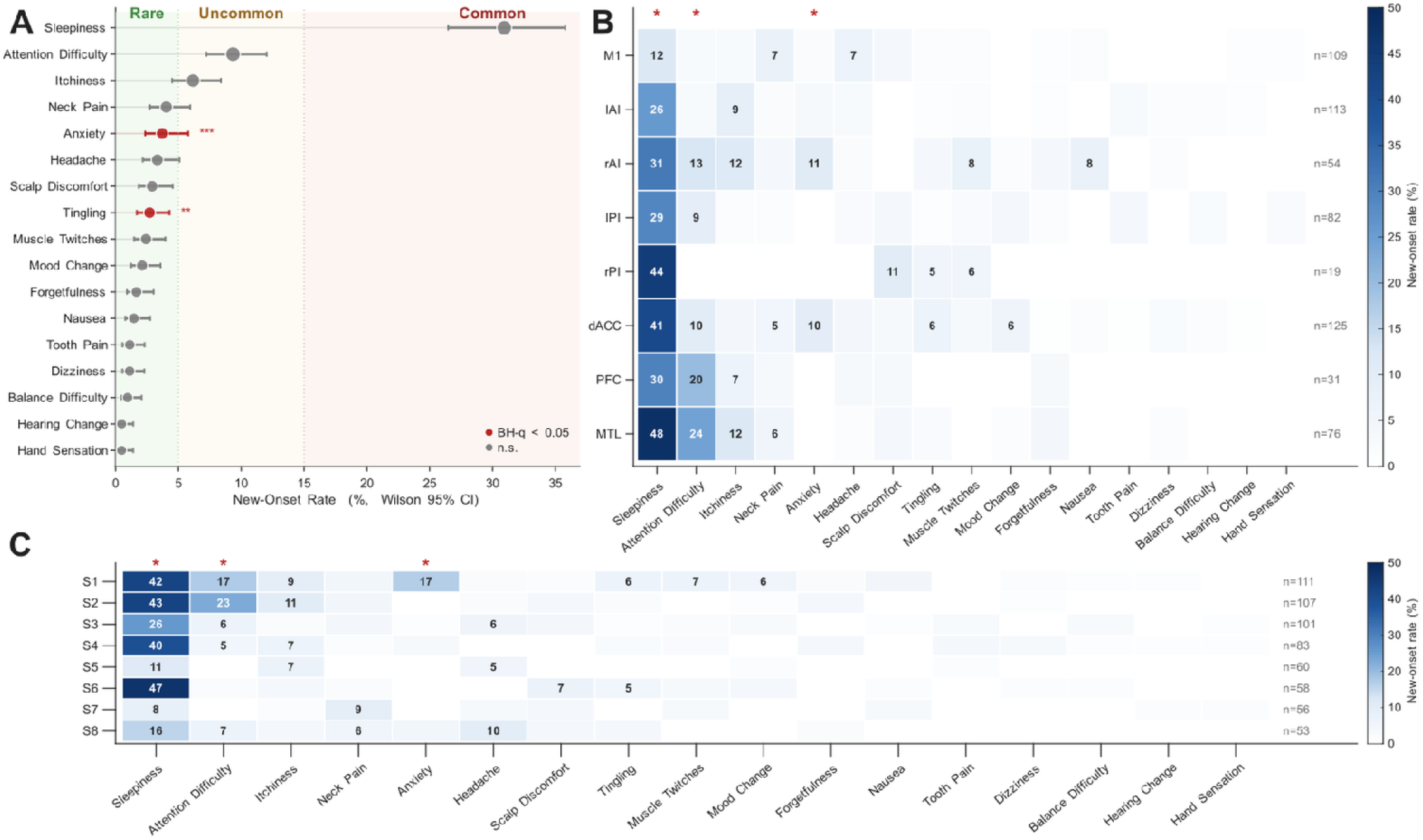
New-onset symptom incidence across all 629 sessions, resolved by symptom domains, brain target, and study. New-onset events were defined as a symptom rated mild or greater at the post-intervention assessment among sessions in which that symptom was absent at pre-intervention baseline. The at-risk denominator for each rate includes only sessions free of that symptom at baseline. Rates are pooled across all 629 sessions. **A.** New-onset rate (%) with Wilson 95% confidence intervals for all 17 symptom domains, ordered from lowest to highest rate (bottom to top). Each domain is plotted as a filled circle whose size scales with the number of new-onset events contributing to that rate. Background shading delineates three frequency zones, rare (0–5%, green), uncommon (5–15%, yellow), and common (>15%, red). The circle and its confidence interval are colored red where the overall pre-to-post change in endorsement for that domain was significant (McNemar test, BH-q correction). Adjacent significance markers denote the level (* BH-q < 0.05, ** BH-q < 0.01, *** BH-q < 0.001). **B.** New-onset rate (%) for each of the eight brain targets (rows, M1, lAI, rAI, lPI, rPI, dACC, PFC, MTL) across all 17 symptom domains (columns). Columns are ordered by overall new-onset rate from highest to lowest (left to right). Cell color denotes new-onset rate (0 to 50%), with darker colors indicating higher new-onset rates. The rate is printed within cells at or above 5%, and gray cells indicat fewer than three at-risk sessions. The session count per target is shown at right. Red asterisks above a column denote which new-onset rate differed across targets (label-permutation χ² test, 10,000 permutations, B -q correction across the 17 domains, BH-q < 0.05). The test identifies that a domain varies across targets rather than which specific target differs. The per-cell rates show where the highest and lowest values fall. **C.** New-onset rate (%) for each of the eight studies (rows, S1–S8) across the same 17 symptom domains, plotted on the color scale and column order of panel B. The session count per study is shown at right, and asterisks mark domains for which new-onset rate differed across studies.

New-onset rates varied by brain target, with sleepiness having the highest new-onset rate of any domain at all eight targets (**Figure 3B**). Three domains showed significant variation in new-onset rate across targets (label-permutation χ² test, 10,000 permutations, BH-q correction): sleepiness (12.2% at M1 to 47.9% at MTL, BH-q = 0.006), attention difficulty (3.0% at M1 to 24.3% at MTL, BH-q = 0.002), and anxiety (0.0% at four targets to 10.8% at rAI, BH-q = 0.047). No other domain differed significantly across brain targets (**Figure 3B**).

New-onset rates also varied by study, with sleepiness ranging from 8.3% in S7 to 47.1% in S6 (**Figure 3C**). Three domains showed significant variation in new-onset rate across studies (label-permutation χ² test, 10,000 permutations, BH-q correction): sleepiness (BH-q < 0.001), attention difficulty (0.0% in S5 and S7 to 23.0% in S2, BH-q < 0.001), and anxiety (0.0% in four studies to 16.9% in S1, BH-q < 0.001). No other domain differed significantly across studies (**Figure 3C**).

### Symptom Severity Transitions

*Anxiety and Sleepiness.* Anxiety and sleepiness were examined in detail as the two most commonly endorsed domains at baseline (sleepiness 40.4%, anxiety 22.7%) and the two domains with a significant within-session change at the full-dataset level, anxiety in endorsement (McNemar BH-q = 1.08 × 10⁻□) and sleepiness in severity (Wilcoxon signed-rank BH-q = 0.007). Severity transitions for the remaining 15 domains are shown in **Supplementary Figure S1**.

Anxiety severity decreased after intervention (Wilcoxon signed-rank BH-q < 0.001, n = 629, **Figure 4A**), driven by 71 sessions that moved from mild to absent against 18 that moved from absent to mild, moderate, or severe (**Figure 4B**). Post-intervention anxiety was endorsed in 11.7% of LIFU and 19.7% of sham sessions (Fisher p = 0.015, **Figure 4C**).

**Figure 4.**
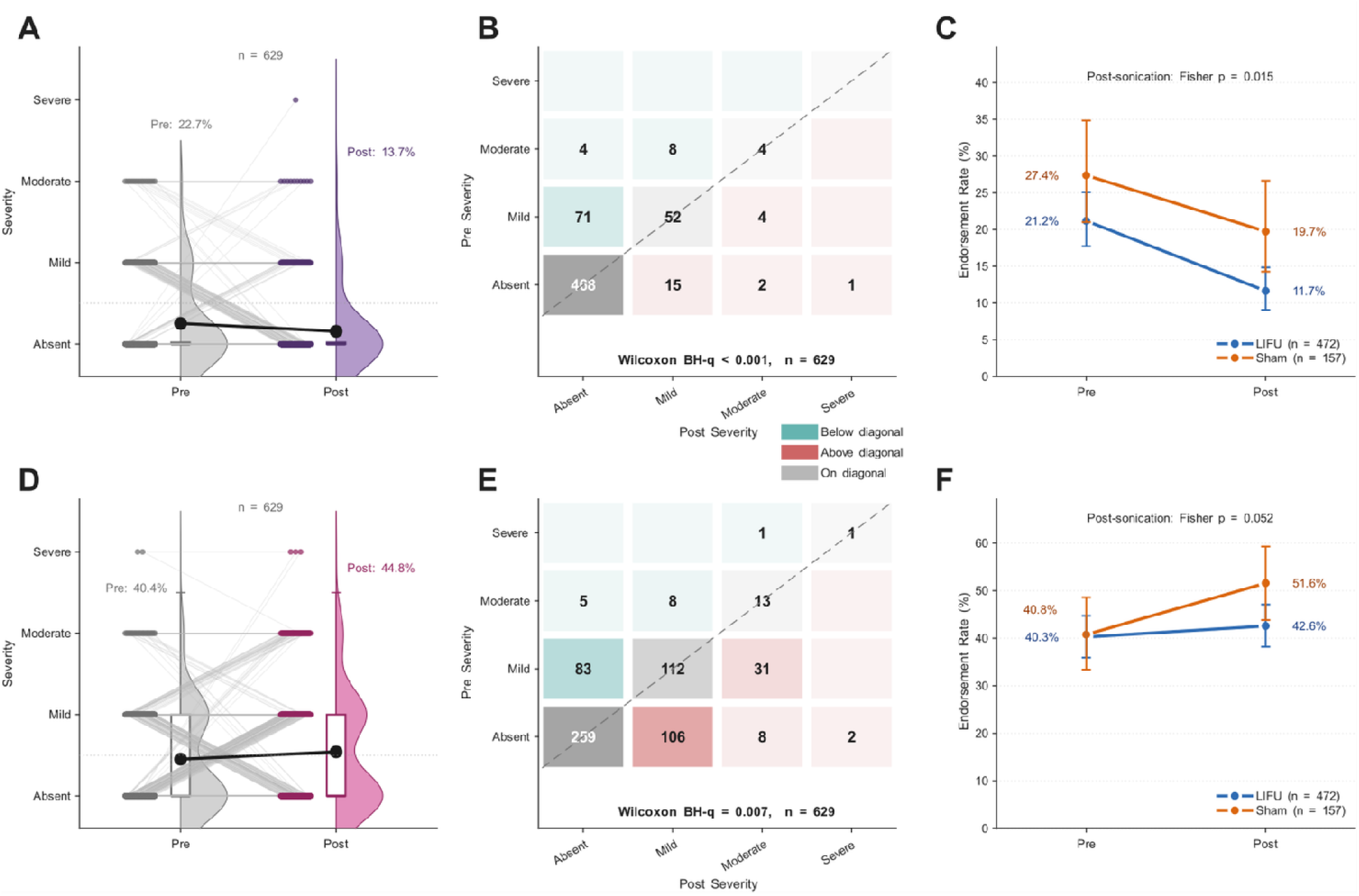
Within-session severity transitions and LIFU-versus-sham endorsement for anxiety and sleepiness. **A.** Raincloud plot of anxiety severity (absent, mild, moderate, severe) at the pre-intervention (gray) and post-intervention (purple) assessments across all 629 sessions. The half-violin shows the kernel density estimate of the severity distribution, the box plot marks the median and interquartile range (IQR) with whiskers extending to 1.5 × IQR, individual session values are overlaid as stacked dots, and gray lines connect the paired pre- and post-intervention values within each session. Black circles with error bars denote the group mean plus or minus the standard error of the mean (SEM). **B.** Anxiety severity-transition matrix in which each cell is the number of sessions moving from a pre-intervention severity category (rows) to a post-intervention severity category (columns). Teal cells above the diagonal mark sessions whose severity decreased, red cells below the diagonal mark sessions whose severity increased, gray cells on the diagonal mark unchanged sessions, and cell opacity scales with the session count. The Wilcoxon signed-rank BH-q for the severity change and the number of sessions are annotated below the matrix. **C.** Anxiety endorsement rate at the pre-and post-intervention timepoints, plotted separately for LIFU (blue) and sham (orange) sessions, with each point showing the endorsement rate and its Wilson 95% confidence interval. The Fisher exact p-value comparing the post-intervention endorsement rates of the two conditions is annotated within the panel. **D.** Raincloud plot of sleepiness severity, with the post-intervention distribution shown in pink, formatted as in A. **E.** Sleepiness severity-transition matrix, formatted as in B. **F.** Sleepiness endorsement rate for LIFU (blue) and sham (orange) sessions, formatted as in C. BH-q denotes the Benjamini-Hochberg false discovery rate adjusted p-value.

Sleepiness endorsement was 40.4% before and 44.8% after intervention (McNemar BH-q = 0.20, **Figure 2**), but severity increased across the ordinal scale (Wilcoxon signed-rank BH-q = 0.007, n = 629, **Figure 4D**), driven by 106 sessions that moved from absent to mild and 10 that reached moderate or severe (**Figure 4E**). Post-intervention sleepiness was endorsed in 42.6% of LIFU and 51.6% of sham sessions (Fisher p = 0.052, F**igure 4F**).

### LIFU versus sham Comparison

No domain showed a statistically significant difference in new-onset rate between LIFU and sham sessions after BH-q correction across all 17 domains (n = 472 LIFU and 157 sham, Fisher’s exact test, smallest BH-q = 0.16). Of the 17 domains, three were nominally higher, but non-significant, in sham than in LIFU: sleepiness (sham 39.8%, LIFU 28.0%, Fisher p = 0.039), itchiness (sham 11.0%, LIFU 4.6%, Fisher p = 0.009), and mood change (sham 4.6%, LIFU 1.3%, Fisher p = 0.022), none of which survived BH-q correction (all BH-q ≥ 0.16) (**Figure 5A**). Headache was the only domain nominally higher, but non-significant, in LIFU compared to sham (LIFU 4.2%, sham 0.7%, Fisher p = 0.059, BH-q = 0.25) (**Figure 5A**).

**Figure 5.**
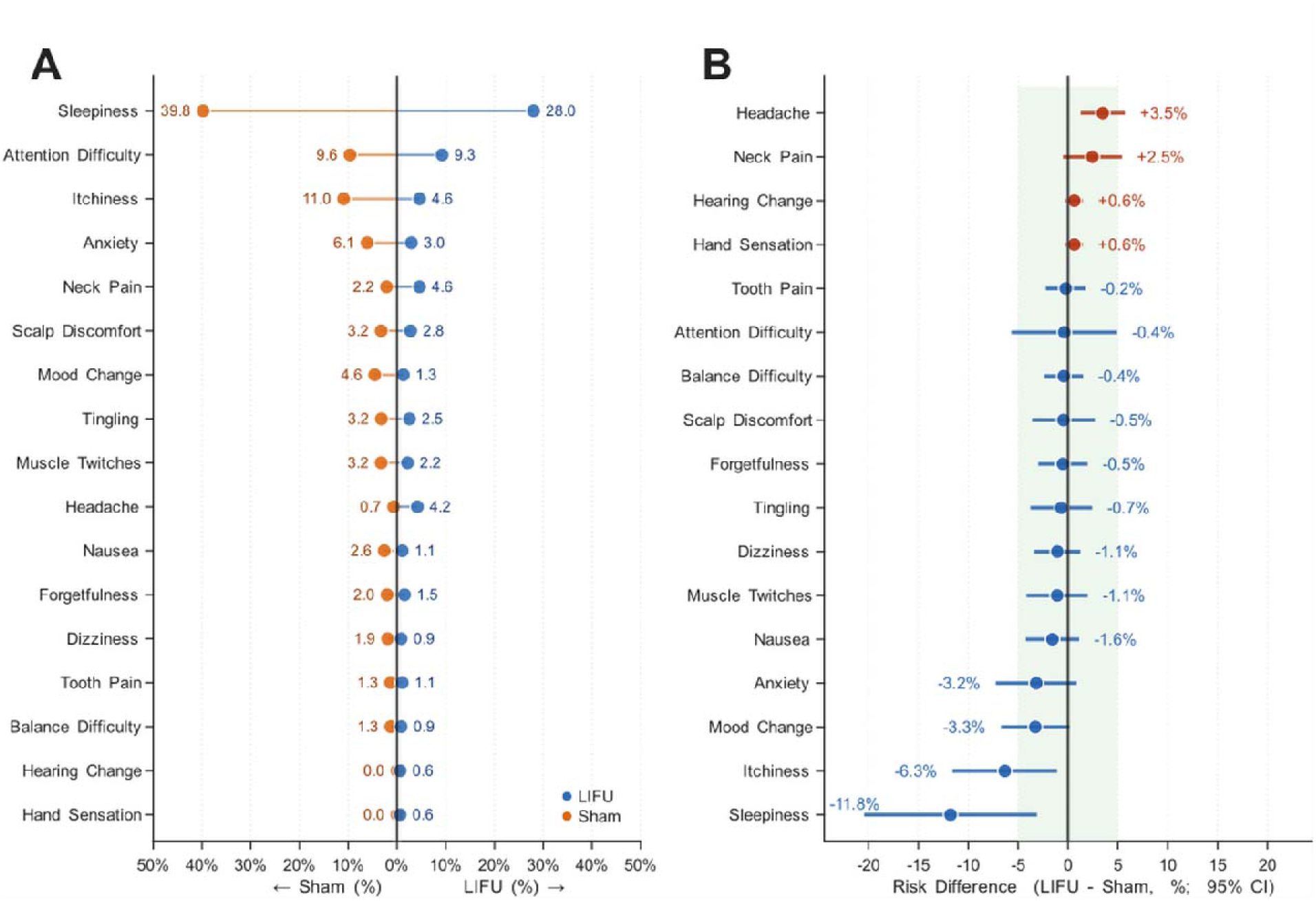
New-onset symptom rates and LIFU-sham risk differences across 17 Report of Symptoms (ROS) domains. New-onset was defined as a symptom rated mild or greater at the post-intervention assessment that was absent at pre-intervention baseline, and the new-onset rate is the percentage of at-risk sessions (those free of that symptom at baseline) in which the symptom newly appeared. **A.** Back-to-back lollipop plot of new-onset rates for LIFU (blue, right) and sham (orange, left) sessions (n = 472 and 157, respectively), with symptoms sorted in descending order by mean new-onset rate. Values adjacent to each dot indicate the new-onset rate (%) for LIFU and for sham. **B.** Absolute risk difference (ARD) per symptom domain, defined as the LIFU new-onset rate minus the sham new-onset rate (in percent), with 95% confidence intervals, sorted in ascending order by ARD. Dots are colored red where the LIFU rate exceeded the sham rate and blue where the sham rate was equal to or greater. The shaded green region marks the ±5% equivalence margin. A domain whose ARD and full 95% confidence interval fall within this band is deemed equivalent between conditions, whereas an interval extending beyond the band marks a difference that cannot be excluded.

Absolute risk differences (ARDs) were calculated as the LIFU new-onset rate minus the sham new-onset rate, where positive values indicate higher rates in LIFU and negative values indicate higher rates in sham. ARDs ranged from −11.8% (sleepiness) to +3.5% (headache), with 95% confidence intervals crossing zero for 14 of 17 domains, indicating no statistically reliable difference in new-onset rate between LIFU and sham for those domains (**Figure 5B**). Fifteen of 17 domains had ARD point estimates within a ±5 percentage-point band around zero, the range within which a between-condition difference is considered too small to be meaningful. Sleepiness (−11.8%) and itchiness (−6.3%) were the only exceptions, both reflecting nominally higher, but non-significant, rates in sham (**Figure 5B**).

### Total Symptom Burden

Total symptom burden was quantified as the number of ROS domains endorsed within each session, a count ranging from 0 to 17. Participants endorsed approximately one symptom per session on average, and this burden did not increase following LIFU or sham intervention in either condition (**Figure 6A, B**). In LIFU sessions (n = 472), the mean number of endorsed domains was 0.94 (SD 1.10, median 1) before intervention and 1.03 (SD 1.23, median 1) after, a change that was not statistically significant (Wilcoxon signed-rank W = 13528, p = 0.120). Sham sessions (n = 157) followed the same pattern, with 1.29 endorsed domains (SD 1.52, median 1) before and 1.37 (SD 1.45, median 1) after (W = 1948, p = 0.557). Burden remained low across the dataset: post-intervention, 89.4% of LIFU sessions and 85.4% of sham sessions had two or fewer domains endorsed, 43.0% of LIFU sessions had none, and no session endorsed more than seven of the 17 domains.

**Figure 6.**
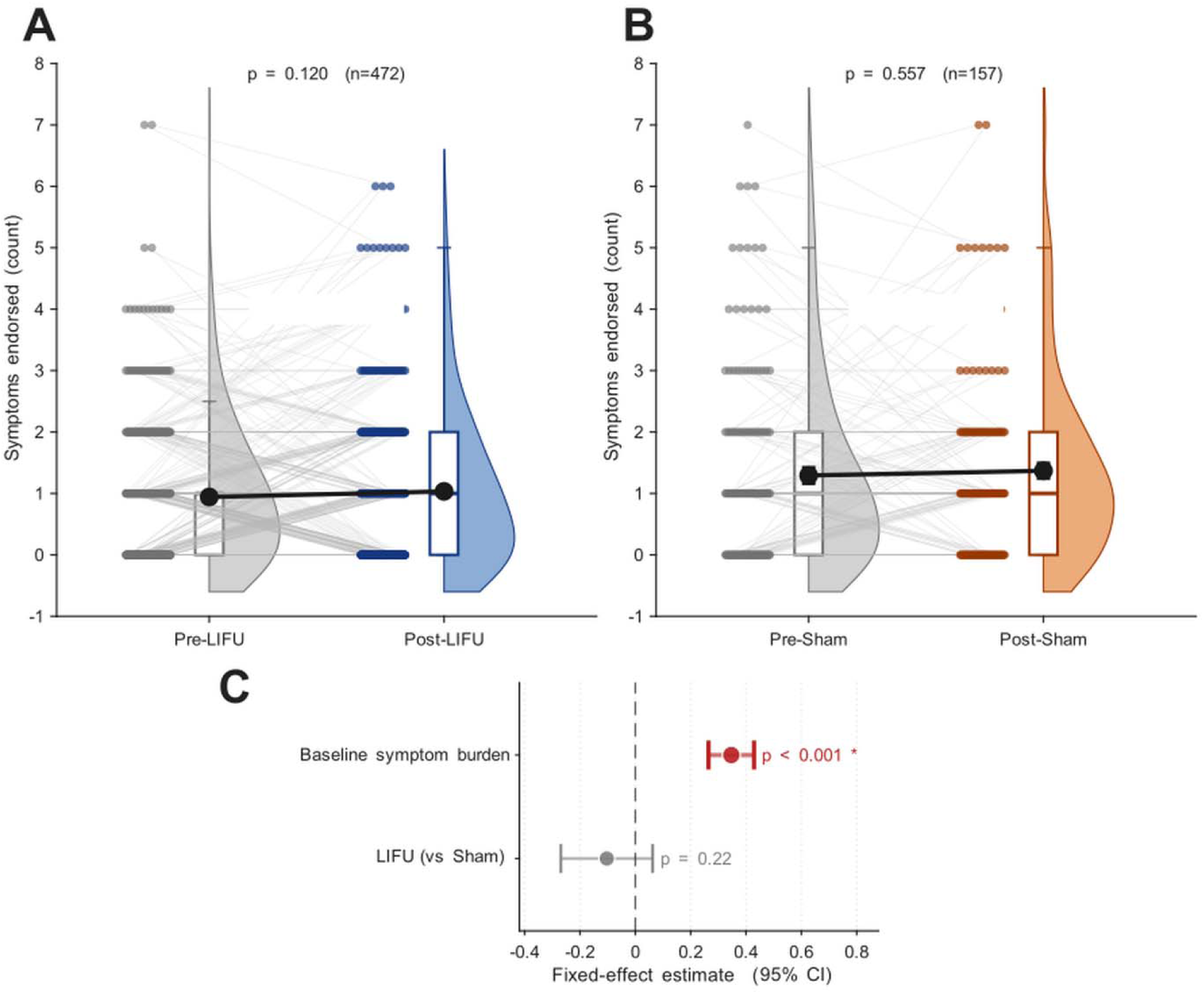
Total per-session symptom burden before and after intervention, and predictors of post-intervention burden. Total symptom burden was defined as the number of Report of Symptoms (ROS) domains endorsed (rated mild or greater) within a session, an integer count from 0 to 17. **A.** Total symptom burden at the pre-LIFU and post-LIFU assessments of each LIFU session (n = 472), shown as vertical raincloud plots. The half-violin shows the kernel density estimate of the symptom-count distribution. The embedded box plot marks the median and interquartile range (IQR), with whiskers extending to 1.5 × IQR. Individual session values are overlaid as stacked dots, and gray lines connect the paired pre- and post-intervention values within each session. Black circles with error bars denote the group mean ± standard error of the mean (SEM). The Wilcoxon signed-rank p-value and session count are annotated within the panel. **B.** Total symptom burden at the pre-sham and post-sham assessments of each sham session (n = 157). **C.** Fixed-effect coefficient estimates from a linear mixed-effects model predicting post-intervention total symptom burden, with participant as a random intercept. Predictors are baseline (pre-intervention) symptom burden and stimulation condition (LIFU versus sham). Filled circles denote point estimates (β) and horizontal lines the 95% confidence intervals. Red marks a statistically significant predictor and gray a non-significant predictor. The dashed vertical line indicates the null value (β = 0).

### Predictors of Post-Intervention Symptom Burden

A linear mixed-effects model tested two predictors of post-intervention total symptom burden, baseline (pre-intervention) symptom burden and stimulation condition (LIFU versus sham), with participant as a random intercept. Only baseline burden predicted post-intervention burden (β = 0.347, SE = 0.042, p < 0.001), so a participant’s post-intervention symptom count was set by their baseline count rather than by the intervention (**Figure 6C**). Stimulation condition did not predict post-intervention burden (β = −0.103, SE = 0.085, p = 0.222).

### Longitudinal Safety Across Repeated LIFU Exposure

New-onset symptom rates for LIFU sessions (n = 472 at-risk sessions) were examined as a function of within-participant LIFU session number, defined as the chronological order of each participant’s LIFU sessions, to test whether accumulating exposures altered symptom emergence. Participants received between 1 and 27 LIFU sessions each, delivered a median of 19 days apart (IQR 7–52) and spanning up to 3.8 years within a single participant. Across all 472 LIFU sessions, the pooled new-onset rate was 35.4%, reflecting the proportion of at-risk sessions in which at least one ROS domain was newly endorsed at post-intervention. New-onset rates across the eight session-number bins ranged from 26.6% at sessions 6–8 (95% CI 15.6–37.5%) to 49.4% at session 2 (95% CI 38.6–60.2%), and the bootstrapped 95% confidence intervals overlapped across all bins. Within-participant total number of LIFU sessions was not a significant predictor of new-onset probability (GLME β = −0.009, SE = 0.025, OR = 0.99, p = 0.73) (**Figure 7**).

**Figure 7.**
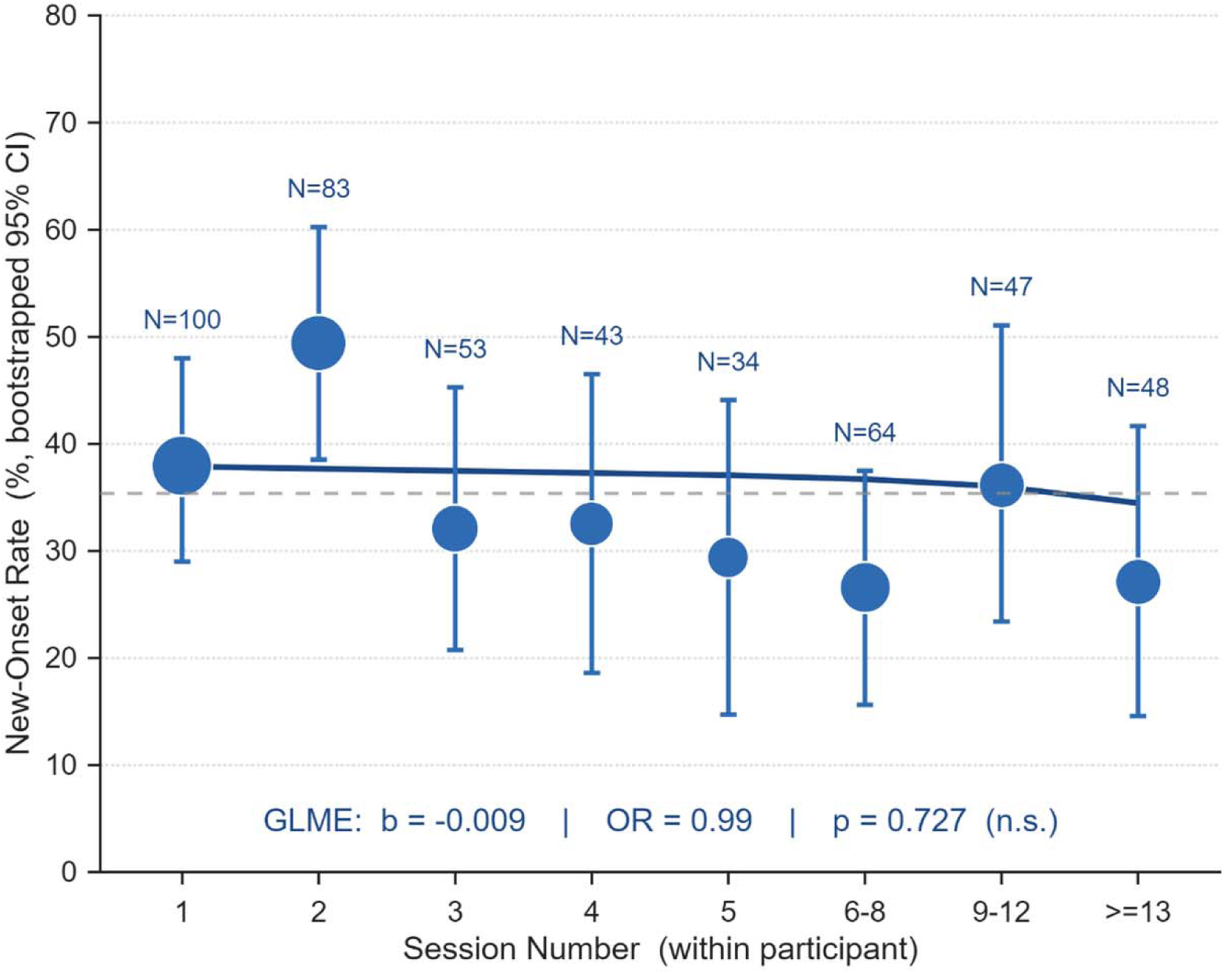
Participation in multiple LIFU sessions does not increase new-onset symptom burden. New-onset rate (%) for LIFU sessions (n = 472) is shown across eight bins of within-participant LIFU session number, the chronological order of each participant’s LIFU sessions (1, 2, 3, 4, 5, 6–8, 9–12, and ≥13). The first five sessions are kept as single-session bins, where nearly every participant contributes, and later sessions are grouped into progressively wider bins (6–8, 9–12, ≥13), where fewer participants contribute, so that each bin retains enough sessions for a stable rate estimate. Filled circles are scaled proportionally to the number of LIFU sessions contributing to each bin, with that count printed above each point. Vertical bars represent bootstrapped 95% confidence intervals (5,000 iterations). The dashed horizontal line marks the pooled LIFU new-onset rate (35.4%). The solid curve depicts the logistic generalized linear mixed-effects model (GLME) trend estimated from fixed effects only (β = −0.009, OR = 0.99, p = 0.73)

### Relationship between Intensity and Symptom Incidence

Across six studies with a sham condition (363 LIFU and 157 sham sessions), extracranial ISPPA ranged from 3.9 to 33.3 W/cm² (**Figure 8A**). New-onset rates in LIFU sessions ranged from 18.6% in S5 to 45.2% in S1. New-onset rate showed a weak, non-significant positive association with extracranial ISPPA (Spearman ρ = 0.37, p = 0.50), and the positive direction was preserved across all leave-one-out iterations (ρ = 0.10 to 0.60). In sham sessions, new-onset rate ranged from 23.5% in S5 to 63.2% in S1 and showed no association with extracranial ISPPA (Spearman ρ = −0.09, p = 0.92, **Figure 8B**).

**Figure 8.**
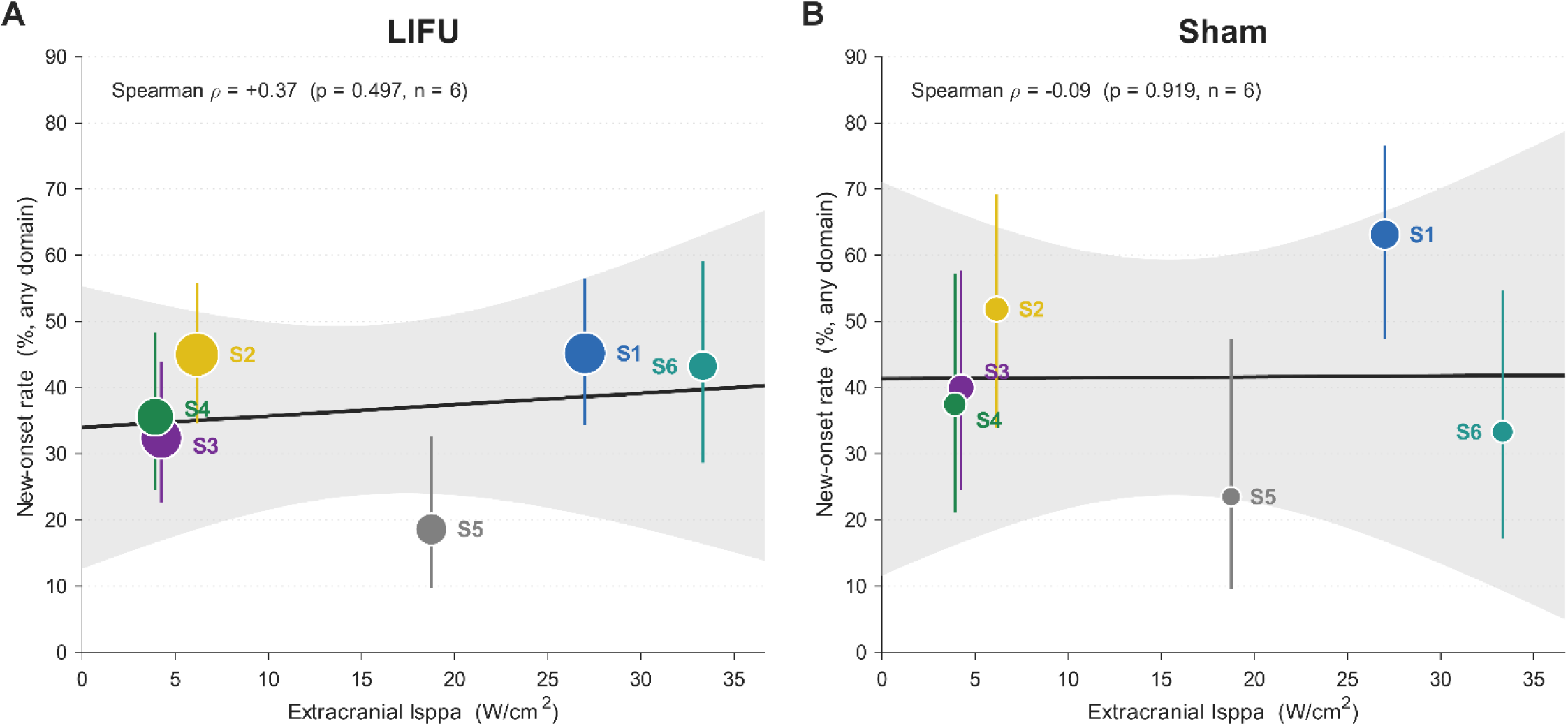
Relationship between extracranial acoustic intensity and new-onset symptom rate, with a sham negative control. Each point is one study, plotted at that study’s extracranial (free-water) spatial-peak pulse-average intensity (ISPPA, W/cm², Table 1) against its new-onset symptom rate. New-onset rate was defined as the proportion of sessions with at least one of the 17 Report of Symptoms (ROS) domains rated mild or greater at the post-intervention assessment that was absent at pre-intervention baseline. Six studies with a sham arm are included (S1, S2, S3, S4, S6, and S5). For S5, which did not report an intensity, ISPPA was calculated from its reported 750 kPa free-water pressure as ISPPA = P²/2ρc, giving 18.75 W/cm². S7 and S8 were excluded because they delivered two intensity levels that cannot be resolved to individual sessions and included no sham sessions. **A.** LIFU sessions (n = 363). Each study is a filled circle whose area scales with the number of LIFU sessions contributing to that study and whose color denotes the study (S1 to S6). Vertical bars are Wilson 95% confidence intervals for the rate. The solid line is the ordinary-least-squares regression of new-onset rate on ISPPA and the shaded band its 95% confidence interval. The Spearman rank correlation and its p-value is annotated within the panel (ρ = +0.37, p = 0.50). **B.** sham sessions (n = 157), plotted identically as a negative control, with each study’s sham sessions assigned that study’s extracranial ISPPA (ρ = −0.09, p = 0.92).

### Symptom Safety in Clinical Pain Populations

Total symptom burden before and after intervention was assessed in two clinical pain cohorts. In Cohort 1 (fibromyalgia, n = 13), total symptom burden did not increase after intervention in either LIFU or sham sessions, moving from 5.46 to 4.38 endorsed domains in LIFU sessions (Wilcoxon signed-rank p = 0.016) and from 5.85 to 5.00 in sham sessions (p = 0.008, **Figure 9A**). In Cohort 2 (chronic pain, n = 22), total symptom burden did not change after intervention in either condition, moving from 4.14 to 3.73 endorsed domains in LIFU sessions (p = 0.458) and from 3.86 to 3.77 in sham sessions (p = 0.684, **Figure 9B**).

**Figure 9.**
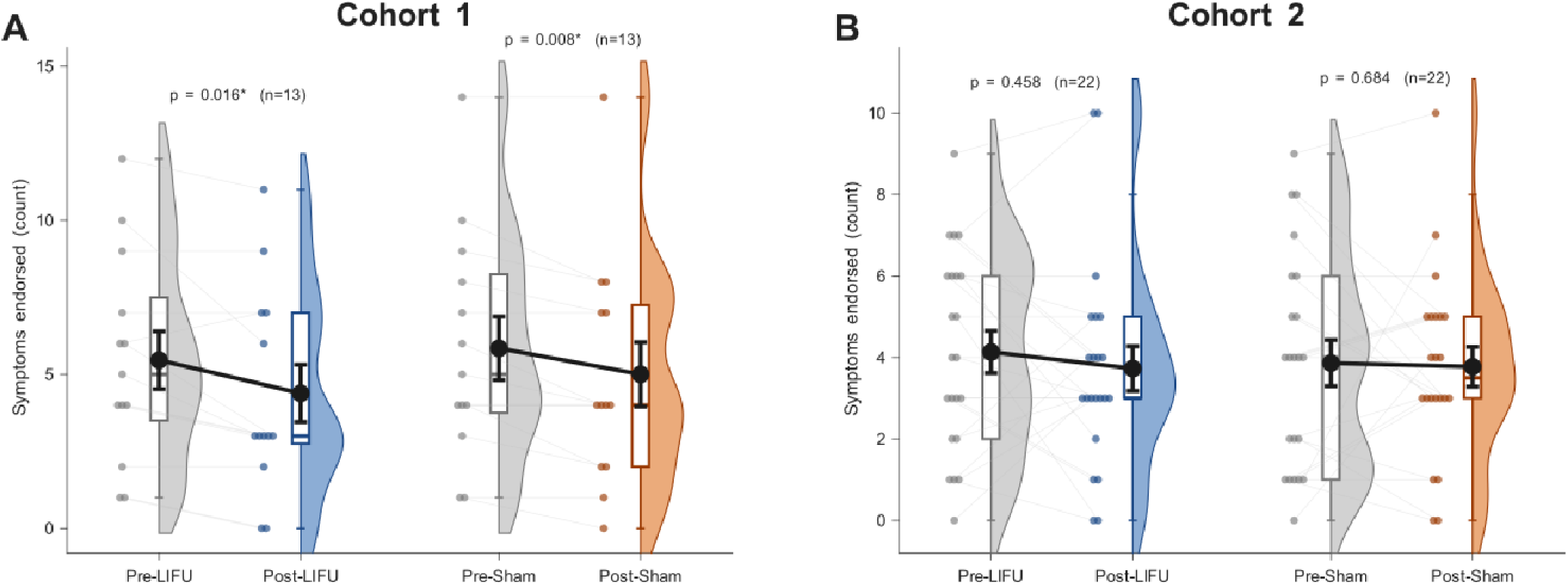
Total symptom burden before and after intervention in two clinical pain populations. Within- session Report of Symptoms (ROS) data are shown for two cohorts analyzed independently of the healthy-adult dataset, Cohort 1 (fibromyalgia) and Cohort 2 (general chronic pain). Total symptom burden was defined as the number of ROS domains endorsed, rated mild or greater, within a session, an integer count from 0 to 17. **A.** Total symptom burden at the pre- and post-intervention assessments of LIFU and sham sessions in Cohort 1, shown as vertical raincloud plots. The half-violin shows the kernel density estimate of the symptom-count distribution. The embedded box plot marks the median and interquartile range (IQR), with whiskers extending to 1.5 × IQR. Individual session values are overlaid as stacked dots, and gray lines connect the paired pre- and post-intervention values within each session. Pre-intervention distributions are shown in gray, the post-LIFU distribution in blue, and the post-sham distribution in orange. Black circles with error bars denote the group mean ± standard error of the mean (SEM). The Wilcoxon signed-rank p-value and session count are annotated within the panel for the LIFU and the sham comparison, where an asterisk marks p < 0.05. **B.** Total symptom burden in Cohort 2, plotted identically to A.

## DISCUSSION

This updated Report of Symptoms dataset comprised 629 sessions of LIFU or sham stimulation delivered to 106 healthy adult volunteers across eight studies and eight cortical and subcortical targets, spanning extracranial ISPPA values of 3.9 to 33.3 W/cm² and mechanical index values of 0.5 to 1.4. A separate clinical sample of 35 patients with chronic pain conditions (13 with fibromyalgia and 22 with heterogeneous chronic pain) was examined independently. Across this range, LIFU produced no serious adverse events and no post-intervention symptom profile distinguishable from sham across all 17 monitored domains. Pre-intervention symptom burden was the only predictor of post-intervention burden, while stimulation condition (LIFU versus sham) contributed no independent effect. Symptom emergence did not increase across repeated exposures within individual participants, and the rate of newly reported symptoms showed only a weak, non-significant relationship with acoustic intensity across studies. These results constitute the largest prospectively collected safety dataset for human LIFU neuromodulation to date and establish that, within established biophysical safety thresholds, LIFU adds no meaningful symptom burden and is well tolerated by healthy adults, with preliminary evidence of the same tolerability in 35 patients with chronic pain conditions.

This work extends our original report [12], which retrospectively compiled ROS data from 120 participants across seven experiments and described a mild, transient symptom profile dominated by sleepiness, headache, and scalp tingling. The present dataset reproduces that profile prospectively, with paired pre- and post-intervention assessment at every session and a sham comparison that the earlier work lacked.

Reproducing it across 629 sessions, eight independent studies, and eight different cortical and subcortical targets establishes that the favorable symptom profile is not specific to a single brain region, study population, or parameter set, but is a general feature of LIFU delivered within current neuromodulation exposure regimes.

The central safety finding is the absence of a symptom signal attributable to LIFU sonication. No symptom domain differed in new-onset rate between LIFU and sham after correction for multiple comparisons. Total symptom burden, expressed as the number of distinct domains a participant endorsed in a session, averaged approximately one symptom per session and did not rise after intervention for either LIFU or sham. Once baseline symptom state was accounted for, the number of symptoms a participant reported after intervention tracked the number they reported beforehand, and whether the session delivered LIFU or sham did not contribute to that prediction. Participant-experienced symptoms in these studies are therefore better understood as reflecting baseline state and the demands of a prolonged experimental visit than the acoustic exposure itself.

Two domains warrant individual attention because each showed a within-session change at the full-dataset level. Anxiety was the most prevalent symptom at baseline after sleepiness and was the only domain whose endorsement rate changed meaningfully from pre- to post-intervention, falling from 22.7% to 13.7%. Because the pre-intervention ROS is administered at the start of the visit, before any intervention, this decline most plausibly reflects the natural attenuation of anticipatory arousal as a participant settles into a familiar procedure rather than any anxiolytic action of LIFU. Sleepiness, the most commonly endorsed domain at baseline and the most frequent new-onset symptom, increased in severity after intervention without a corresponding change in overall incidence. This pattern, a deepening of an already present symptom rather than the appearance of a new one, is consistent with accumulating fatigue over an experimental session, as LIFU and sham sessions did not differ. Tingling was the only domain with a meaningful net increase in new onset, yet its rate was equivalent in LIFU and sham sessions, consistent with a transducer-to-scalp contact mechanism shared by both conditions and with prior work showing that participants do not reliably distinguish LIFU from sham coupling in masked LIFU designs [19,23].

A question left largely unanswered by single-condition symptom counts is whether the intensity of LIFU sonication shapes the symptoms participants report. Our original report observed a positive correlation between spatial-peak pulse-average intensity and symptom response rate across experiments, and the present dataset allowed a prospective re-examination of that relationship across the six studies that delivered a defined extracranial ISPPA alongside a sham condition. The rate of newly reported symptoms in LIFU sessions increased with extracranial ISPPA (3.9 to 33.3 W/cm²), whereas the same rate in sham sessions showed no such relationship. This gradient did not reach statistical significance, and its positive direction was preserved when any single study was removed, indicating a consistent but modest trend rather than a definitive dose-response effect. Two features temper its interpretation. The unit of analysis was the study rather than the session, because intensity was fixed within a protocol and could not be attributed to individual sessions, which limits the comparison to a handful of points. Extracranial ISPPA also indexes intensity in free water rather than the pressure ultimately delivered to the brain, which varies with skull and beam geometry within and between participants. The convergence of a positive intensity-symptom relationship in LIFU but not sham sessions, replicating the direction reported previously [12], nonetheless points to a graded contribution of acoustic exposure to tolerability. Resolving it will require session-level estimates of the intracranial exposure, modeled per participant and standardized across devices, so that exposure can be related to symptoms at the level of the individual session rather than the study [16].

As LIFU advances toward therapeutic use, where treatment will require many sessions rather than one, the safety of repeated exposure within an individual becomes a central question that single-session reports cannot address. The multi-study structure of this dataset offered an initial preliminary examination as a subset of participants enrolled across several protocols and accumulated up to 27 LIFU sessions each, delivered a median of 19 days apart and spanning as long as 3.8 years within one participant. The rate at which new symptoms emerged did not increase as participants accumulated sessions, fluctuating within a narrow band with overlapping confidence intervals across the full range of exposure. This stability is consistent with evidence that focused ultrasound at neuromodulatory parameters does not produce cumulative tissue effects [4,7,29,30] and provides direct, within-person data that repeated LIFU does not progressively increase symptom burden. For investigators designing multi-session therapeutic protocols, these data offer an early empirical basis for delivering LIFU repeatedly to the same individual without an expectation of escalating participant-experienced symptoms. These sessions were spaced over days to weeks, however, whereas many therapeutic protocols deliver LIFU in closer succession, sometimes several times within a single day. The present data therefore speak to repeated exposure across spaced sessions and cannot be assumed to extend to within-day or otherwise condensed treatment schedules.

This profile is not limited to healthy volunteers. When the same ROS was applied in two clinical pain cohorts, fibromyalgia and chronic pain, total symptom burden did not rise after intervention in either cohort, and no pattern separated LIFU from sham, despite the substantial baseline symptom burden these patients carry. The cohorts are small and were not designed for formal safety inference, so they cannot establish clinical safety on their own. They do show, however, that the within-session tolerability observed in healthy adults persists in patients with pre-existing symptoms, the setting in which LIFU is most likely to be applied therapeutically.

These findings sit within a broader literature that has consistently found no evidence of structural brain injury from LIFU at neuromodulatory exposures [10]. They acquire particular relevance given a recent serious adverse event in which focused ultrasound delivered to the nucleus accumbens at acoustic exposures well above those used in any study reported here produced edema, microhemorrhage, and persistent neurological sequelae [13]. The present cohort, drawn entirely from a parameter space consistent with established mechanical and thermal safety envelopes [4,15], showed no analogous signal, a contrast that underscores how strongly participant risk depends on where a protocol sits within the exposure spectrum. That same event prompted the field to discuss the distinction between low-, intermediate-, and high-intensity regimes and to call for standardized reporting of acoustic parameters and adverse events [14,16]. Realizing that goal for participant-experienced symptoms requires a complementary instrument. Published LIFU symptom data are drawn from heterogeneous questionnaires applied at variable timepoints, so differences in reported rates across studies cannot be separated from differences in how symptoms were measured. A symptom framework administered under standardized conditions, as the ROS was here, removes that confound and becomes a prerequisite for meaningful cross-study surveillance as LIFU reaches larger and more varied clinical populations [17].

Non-invasive brain stimulation offers a clear precedent for this trajectory. Consensus safety frameworks for transcranial magnetic stimulation [33] and transcranial direct current stimulation [35] established shared adverse event classifications that later enabled meta-analytic synthesis across hundreds of independently conducted studies [36,37]. LIFU neuromodulation is at an equivalent juncture, with a growing base of clinical trials [38–40], the ITRUSST consensus documents [4,15], and the present dataset together providing sufficient grounds to support field-wide reporting standards. Future studies should pair a standardized participant-reported symptom instrument with the ITRUSST reporting framework, assess symptoms at consistent pre- and post-intervention timepoints, and deposit anonymized symptom data alongside acoustic exposure parameters so that safety evidence can accumulate across laboratories as therapeutic applications expand.

These findings extend the profile first described in 2020 [12] into a prospective, sham-controlled reference for human LIFU neuromodulation, and the initial clinical cohorts indicate that the profile is not confined to healthy volunteers. As LIFU moves further into therapeutic trials, this benchmark, paired with standardized symptom reporting, provides the foundation on which clinical safety monitoring can be built.

## Data availability

The datasets generated and analyzed during the current study are not publicly available to protect participant privacy but are available from the corresponding author on reasonable request.

## Author contributions

A.K. performed data analysis and wrote the manuscript. T.C. compiled the dataset and assisted with manuscript preparation. W.L. conceived the study, supervised analysis, provided resources and funding, and reviewed/edited the manuscript.

