## Supplemental Text and Figures for "An updated prospective quantitative analysis of symptoms and safety in low-intensity focused ultrasound neuromodulation"

**Supplementary Methods**

*LIFU Transducers and Waveforms*. LIFU was delivered using one of three different 500 kHz focused ultrasound transducers selected on a study-by-study basis to accommodate scalp-to-target depth across the brain regions and participants in each protocol. These included: an H-281 (single-element; active diameter 45.0 mm; geometric focus 45.0 mm; focal depth from exit plane 38.0 mm); an H-104 (single-element; active diameter 64.0 mm; geometric focus 63.2 mm; focal depth from exit plane 52.0 mm) and a CTX-500 dual-element annular array; active diameter 60.0 mm; geometric focus 64.0 mm; electronically adjustable focal depth (Sonic Concepts, Bothell, WA, USA).

Transducers were acoustically coupled to the participant's scalp by first applying ultrasound gel directly to the scalp to separate the hair, expose the skin surface, and minimize air at the skin–transducer interface. A custom-fabricated coupling puck was then positioned between the transducer face and the scalp [18,19]. Sonication parameters for each study are reported in **Table 1**.

**Supplementary Figures**

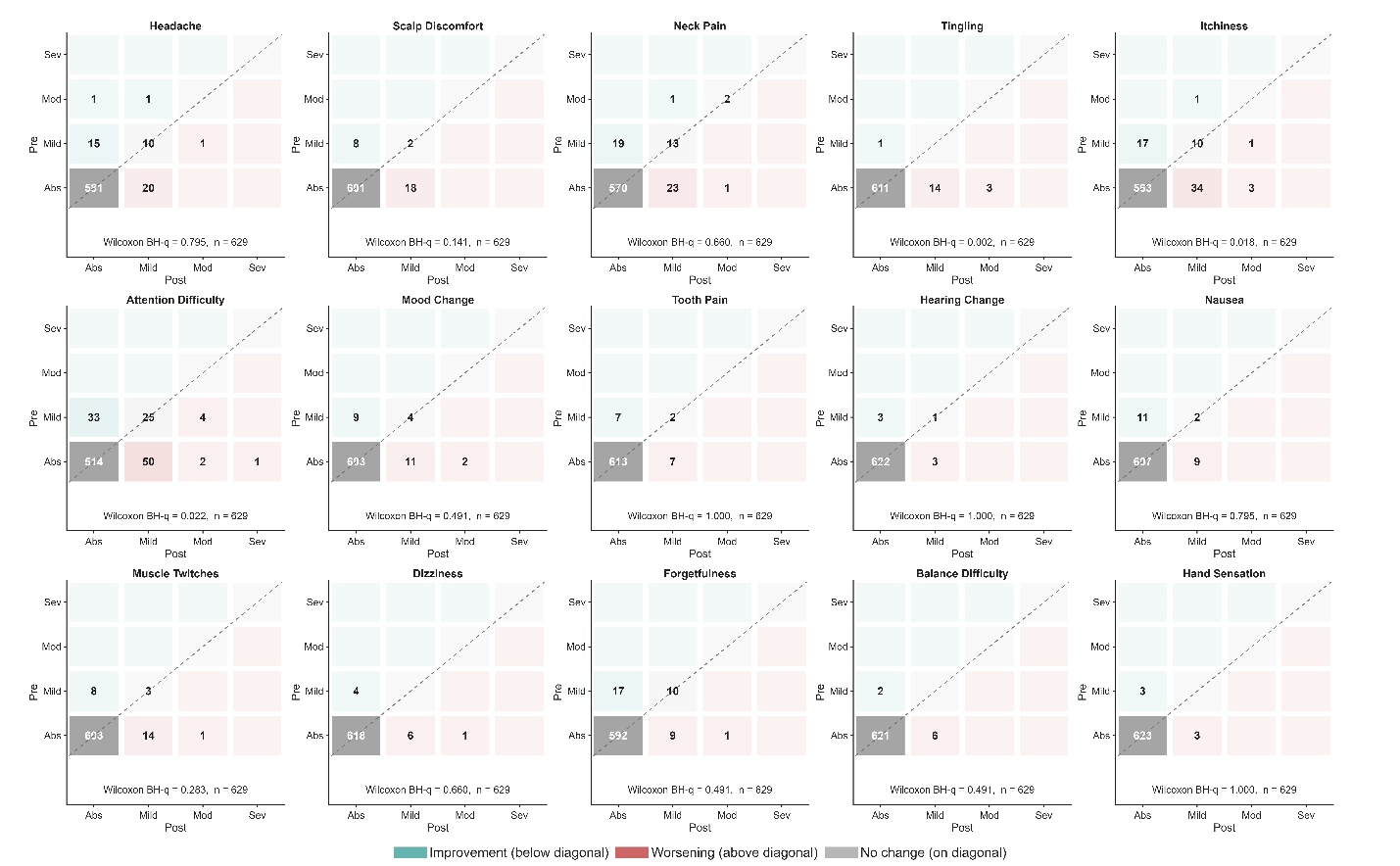

**Supplementary Figure S1. Within-session severity transitions for 15 Report of Symptoms (ROS) domains.** Each panel is a severity-transition matrix for one domain, pooled across all 629 sessions, in which each cell is the number of sessions moving from a pre-intervention severity category (rows: absent, mild, moderate, severe) to a post-intervention severity category (columns). Teal cells above the diagonal mark sessions whose severity decreased, red cells below the diagonal mark sessions whose severity increased, gray cells on the diagonal mark unchanged sessions, and cell opacity scales with the session count. Each panel is annotated with the Wilcoxon signed-rank BH-q for the severity change and the number of sessions with complete pre- and post-sonication ratings (n).
